# Efficacy of remdesivir in hospitalised COVID-19 patients in Japan: A large observational study using the COVID-19 Registry Japan

**DOI:** 10.1101/2021.03.09.21253183

**Authors:** Shinya Tsuzuki, Kayoko Hayakawa, Yukari Uemura, Tomohiro Shinozaki, Nobuaki Matsunaga, Mari Terada, Setsuko Suzuki, Hiroshi Ohtsu, Yusuke Asai, Koji Kitajima, Sho Saito, Gen Yamada, Taro Shibata, Masashi Kondo, Kazuo Izumi, Masayuki Hojo, Tetsuya Mizoue, Kazuhisa Yokota, Fukumi Nakamura-Uchiyama, Fumitake Saito, Wataru Sugiura, Norio Ohmagari

## Abstract

**Objectives:** Although several randomised controlled trials have compared the efficacy of remdesivir with that of placebo, there is limited evidence regarding its effect in the early stage of nonsevere COVID-19 cases.

**Methods:** We evaluated the efficacy of remdesivir on the early stage of nonsevere COVID-19 using the COVID-19 Registry Japan, a nationwide registry of hospitalised COVID-19 patients in Japan. Two regimens (start remdesivir therapy within 4 days from admission vs. no remdesivir during hospitalisation) among patients without the need for supplementary oxygen therapy were compared by a three-step processing (cloning, censoring, and weighting) method. The primary outcome was supplementary oxygen requirement during hospitalisation. Secondary outcomes were 30-day fatality risk and risk of invasive mechanical ventilation or extracorporeal membrane oxygenation (IMV/ECMO).

**Results:** The data of 12,657 cases met our inclusion criteria. The ‘start remdesivir’ regimen showed a lower risk of supplementary oxygen requirement (hazard ratio: 0.861, *p* < 0.001). Both 30-day fatality risk and risk of IMV/ECMO introduction were not significantly different between the two regimens (hazard ratios: 1.05 and 0.886, *p* values: 0.070 and 0.440, respectively).

**Conclusions:** Remdesivir might reduce the risk of oxygen requirement during hospitalisation in the early stage of COVID-19; however, it had no positive effect on the clinical outcome and reduction of IMV/ECMO requirement.

## Introduction

As in other parts of the world, the number of COVID-19 patients is increasing in Japan, with 1,069,554 cases and 15,330 deaths being reported from January 14, 2020, to August 12, 2021[1]. In addition to the treatment of the hyperinflammatory state and coagulopathy, antiviral medication is one of the important components of COVID-19 treatment[2]. Among the antiviral medications for SARS-CoV-2, only remdesivir was approved in Japan on May 7, 2020[3].

Several randomised controlled trials (RCTs) have compared the efficacy of remdesivir with that of placebo. In an RCT in China that enrolled hospitalised COVID-19 pneumonia patients with hypoxia, no statistically significant clinical benefits were observed[4]. A multinational RCT (ACTT-1) conducted in Europe, the United States, and Asia including Japan confirmed that remdesivir shortened the time to recovery in hospitalised COVID-19 patients with pneumonia[5]. However, in the subgroup analysis, no reduction in the time to recovery was observed in patients who were intubated or on extracorporeal membrane oxygenation (ECMO) at the time of drug administration. Although the recovery rate improvement observed among patients enrolled from Asia was similar to that among the overall population, ethnically Asian patients did not show such treatment benefit. The multinational SOLIDARITY Trial, organised by the World Health Organization, demonstrated no survival benefit for remdesivir in hospitalised COVID-19 patients[6]. The trial enrolled 61% of patients from Asia and Africa in total, but no patients from Japan were enrolled. In another RCT conducted in the United States, Europe, and Asia including 16%–19% Asians, No statistically significant difference was observed between the 10-day remdesivir group and the standard treatment group[7]. These data indicate conflicting results regarding remdesivir’s clinical efficacy, and currently, recommendations in the guidelines of remdesivir use against COVID-19 are inconsistent and its optimal role remains uncertain[8].

Although the efficacy of remdesivir against severe COVID-19 cases has been already examined in several studies, its efficacy against nonsevere cases or cases in the early stage of disease has not yet been evaluated. In the study of Spinner et al.[7] targeting patients with COVID-19 pneumonia with preserved room-air oxygen saturation, however, the interpretation of the results of this trial is limited by the inconsistent evidence of the treatment regimens.

We conducted this study to evaluate the efficacy of remdesivir in nonsevere COVID-19 patients in the early stage of disease, especially the stage before the initiation of supplementary oxygen therapy and other pharmaceutical treatment.

## Methods

### Study population and data

We used the data of patients derived from COVIREGI-JP[9]. The inclusion criteria are both (1) a positive SARS-CoV-2 test and (2) inpatient treatment at a healthcare facility. SARS-CoV-2 testing is based on the notification criteria of the Infectious Diseases Law.[10]. Patients who refused to participate in the study by opting out were excluded.

We had modified a case report form (CRF) of the International Severe Acute Respiratory and Emerging Infection Consortium (ISARIC)[11]. Study data were collected and managed using REDCap (Research Electronic Data Capture), Associates and Clinicians (JCRAC) data center of the National Center for Global Health and Medicine.

We used data from cases that had entered all the following major items as of April 30, 2021 (i.e., frozen data as of April 30, 2021), for the present study, similar to the previous report[9]: basic information at admission (demographics and epidemiological characteristics), comorbidities, signs and symptoms at the time of admission (including conditions at admission), outcome at discharge, supportive care during hospitalisation, history of drug administration during hospitalisation, and complications during hospitalisation.

### Study design

#### Eligibility for analysis set

Among all patients registered as cases of COVIREGI-JP, we excluded non-Japanese and <15-year-old cases to evaluate the efficacy of remdesivir in the Japanese adult cohort. We also excluded patients with severe diseases who had already been initiated on supplementary oxygen therapy during admission and/or admitted more than 4 days before the day of symptom onset to evaluate the efficacy of remdesivir in the early stages of treatment.

#### Endpoints, treatment strategies of interest and follow-up

The primary outcome was oxygen therapy requirement during 30 days of admission. The secondary outcomes were 30-day fatality risk and risk of invasive mechanical ventilation (IMV) or ECMO. We compared the following treatment regimens: Regimen 1, start remdesivir therapy within 4 days from the day of admission for at least 3 days and at the most 15 days without the combination of systemic steroids and some antivirals (tocilizumab, baricitinib, and favipiravir), and Regimen 2, not using remdesivir, other antivirals (tocilizumab, baricitinib, and favipiravir), and systemic steroids during their admission. Other supportive treatments were allowed in both regimens.

Each patient was followed up until 30 admission days, event of interest (initiation of oxygen therapy for primary analysis, death, or initiation of IMV/ECMO within 30 admission days for secondary analysis), and discharge, whichever came first. Furthermore, we required both regimens withhold the initiation of supplementary oxygen therapy for 4 days from admission to evaluate the efficacy of remdesivir among patients without the need for intensive therapy at admission. We excluded patients who were initiated on oxygen therapy within 4 days from admission; the possible time-related biases associated with such exclusion after admission (the start of follow-up)[12,13] were addressed by the novel statistical approach described in the next section.

### Statistical analysis

To compare the abovementioned treatment regimens from time-varying remdesivir treatment data in an unbiased manner, we used the ‘three-step’ method (cloning, censoring, and weighting)[14]. First, we prepared clones (or data copies) of patients to assign them to the two regimens on the person-day basis. We assigned each person to the treatment regimens at admission at which the ‘eligibility’ of enrolment was judged. We assigned patients treated with remdesivir at Day 1 to the ‘start remdesivir’ regimen arm and other patients to both regimens. Assigning a patient to both arms simultaneously is equivalent to having two clones of that patient in the dataset, with each copy assigned to a different arm.

Second, we artificially censored the clones if they deviated from their assigned regimen during the follow-up period. For instance, consider a patient who was initiated on remdesivir between Days 1 and 4; his/her clone assigned to Regimen 2 (‘no remdesivir’) was censored at that time, but the clone assigned to Regimen 1 (‘start remdesivir’) was followed up thereafter. Conversely, for a patient not initiated on remdesivir at Day 5, his/her clone assigned to Regimen 1 was censored at Day 5, but the clone assigned to Regimen 2 was followed up thereafter. In addition, clones were censored at any time when the following conditions were met: (1) supported by supplementary oxygen before 4 days from admission, (2) treated with systemic steroids, (3) treated with tocilizumab, (4) treated with baricitinib, (5) treated with favipiravir, (6) duration of remdesivir treatment shorter than 3 days (patients were censored when they discontinued remdesivir before the 3 days elapsed from treatment initiation), and (7) duration of remdesivir treatment longer than 15 days (patients were censored at 15 days if they continued using remdesivir). Moreover, when we compared the primary outcome (supplementary oxygen requirement), patients were censored at the next day of the beginning day of oxygen administration. Similarly, when we compared the secondary outcomes, IMV/ECMO introduction and death within 30 days from admission were the signs of censoring. Discharged patients were censored from the next day of discharge. We set the duration of observation as 30 days, and all patients were censored after 30 days have elapsed since their admission.

Third, to eliminate selection bias due to the abovementioned artificial censoring, we used the inverse probability of censoring weights[15]. The weights of each person-day were calculated using pooled logistic regression models for being censored, such as age, sex, cardiovascular diseases, chronic respiratory diseases, severe renal diseases (serum creatinine level: ≥3 mg/dl) or dialysis, hypertension, hyperlipidemia, obesity diagnosed by physicians, solid tumour, days from symptom onset to admission, use of corticosteroids, use of anticoagulants (time-independent variables), and National Early Warning Score (NEWS, time-dependent variable)[16]. The models were fitted separately according to regimens and follow-up days. The weights were stabilised according to the regimen-day-specific uncensored probability without covariate and were multiplied until that day of the follow-up. Especially, we had only intermittent data about the clinical course of the patient. In this study, we had information of patients at Days 1, 4, 8, 15, 22 and 29. For example, a patient’s record indicating administered oxygen at Day 8 implies that oxygen support for that patient began between Days 5 and 8 and the exact day is not available. We used NEWS at Day 1 as that of Day 1; NEWS at Day 4 as that of Days 2, 3 and 4; NEWS at Day 8 as that of Days 5, 6, 7, and 8; and NEWS at Day 15 as that of Days 9, 10, 11, 12, 13, 14 and 15, same as NEWS at Days 22 and 29. These possible confounders were selected for their potential association with the outcome of interest based on clinical knowledge and previous studies[17–22].

Finally, the discrete-time hazard ratio of primary and secondary outcomes between two regimens was estimated using weighted pooled logistic regression. As each patient has multiple lines in the dataset (each day, each regimen of the same patient until censored), we used cluster-robust standard errors regarding each patient as a cluster to estimate 95% confidence intervals (CIs). We also estimated cumulative incidence rates under the two regimens by multiplying the weighted probabilities of no-event using the Kaplan-Meier method. The pointwise 95% CIs at each day were based on 2.5 and 97.5 percentiles of 1000 bootstrap estimates. All statistical analyses were conducted using R, version 4.0.5[23].

## Results

The data of 12,657 of 16,747 cases met our inclusion criteria. Table 1 describes the basic characteristics of the included cases. A total of 828 patients were treated with remdesivir, and the treatment duration depended on each facility and physician’s decision. The duration of remdesivir treatment was 5 days in 485 cases (58.6%). The 10-day regimen was completed in 106 cases (12.8%). A total of 115 patients (13.9%) were administered remdesivir for <5 days, and 88 (10.6%) were administered between 6 and 9 days. A total of 27 patients (3.3%) were administered for >10 days. Patients in the case group were older, more frequently male and more severe and fatal.

**Table 1.** Basic characteristics of patients who met the inclusion criteria.

|  | Patients initiated on<br>remdesivir<br>(n = 828) | Patients without<br>remdesivir<br>(n = 11,829) | Total<br>(n = 12,657) |
| --- | --- | --- | --- |
| Age (years) | 68 [56–79] | 51 [33–70] | 52 [34–71] |
| Male | 534 (64.5%) | 6276 (53.1%) | 6810 (53.8%) |
| Cardiovascular disease | 57 (6.9%) | 547 (4.6%) | 604 (4.8%) |
| Respiratory disease | 35 (4.2%) | 268 (2.3%) | 303 (2.4%) |
| Diabetes mellitus | 227 (27.4%) | 1416 (12.0%) | 1643 (13.0%) |
| Severe renal disease or dialysis | 19 (2.3%) | 200 (1.7%) | 219 (1.7%) |
| Hypertension | 370 (44.7%) | 2845 (24.1%) | 3215 (25.4%) |
| Obesity | 91 (11.0%) | 819 (6.9%) | 910 (7.2%) |
| Charlson Comorbidity Index | 1 [0–2] | 0 [0–1] | 0 [0–1] |
| NEWS at Day 1 | 1 [0–2] | 1 [0–2] | 1 [0–2] |
| NEWS at Day 4 | 2 [1–4] | 1 [0–2] | 1 [0–2] |
| NEWS at Day 8 | 2 [1–4] | 1 [0–2] | 1 [0–2] |
| NEWS at Day 15 | 2 [1–4] | 1 [0–2] | 1 [0–3] |
| NEWS at Day 22 | 3 [1–5] | 1 [0–3] | 1 [0–3] |
| NEWS at Day 29 | 11 [9–13] | 9 [9–11] | 9 [9–11] |
| Fatal cases | 70 (8.5%) | 285 (2.4%) | 355 (2.8%) |
| Oxygen administration during hospitalisation * | 610 (73.7%) | 1883 (15.9%) | 2493 (19.7%) |
| IMV/ECMO during hospitalisation | 49 (5.9%) | 98 (0.8%) | 147 (1.2%) |
| Days from symptom onset to hospitalisation | 3 [1–4] | 3 [1–4] | 3 [1–4] |
| Use of systemic steroids | 670 (80.9%) | 1840 (15.6%) | 2510 (19.8%) |
| Use of favipiravir | 266 (32.1%) | 2932 (24.8%) | 3198 (25.3%) |
| Use of tocilizumab | 63 (7.6%) | 100 (0.8%) | 163 (1.3%) |
| Use of baricitinib | 0 (0%) | 0 (0%) | 0 (0%) |
| Days from onset to remdesivir administration | 6 [4–9] | NA | NA |
| Days from admission to remdesivir administration | 5 [3–10] | NA | NA |
| Duration of remdesivir administration 5 days | 485 (58.6%) | NA | NA |

Numbers in brackets represent percentage and interquartile range.
NA, not available; NEWS, National Early Warning Score; IMV/ECMO, invasive mechanical ventilation/extracorporeal membrane oxygenation
\* Indication for supplementary oxygen was judged by each physician

Regimen 1 (treated with remdesivir within 4 days of admission) showed a lower risk (adjusted hazard ratio: 0.861, 95% CIs: 0.817–0.907, *p* < 0.001) of supplementary oxygen requirement than Regimen 2 (treated without remdesivir). However, the 30-day fatality risk and risk of IMV/ECMO introduction were not different between the two groups (adjusted hazard ratio: 1.05 [95% CIs: 0.996–1.12] and 0.886 [95% CIs: 0.652–1.20], *p* values: 0.070 and 0.440, respectively). Table 2 shows the details of primary and secondary outcomes.

**Table 2.** Results of pooled logistic regression analysis on the effect of remdesivir on primary and secondary outcomes.

|  | Person-days | Event | Weighted event<br>rate (per 1000<br>person-day) | Hazard<br>ratio | 95% CI | *P* value |
| --- | --- | --- | --- | --- | --- | --- |
| Oxygen<br>requirement |  |  |  |  |  |  |
| Regimen 1 | 84,339 | 223 | 2.61 | 0.861 | 0.817–0.907 | <0.001 |
| Regimen 2 | 84,426 | 258 | 3.03 | 1 | Reference |  |
| 30-day<br>fatality risk |  |  |  |  |  |  |
| Regimen 1 | 86,444 | 33 | 0.394 | 1.05 | 0.996–1.12 | 0.070 |
| Regimen 2 | 86,758 | 33 | 0.379 | 1 | Reference |  |
| IMV/ECMO<sup>*</sup> |  |  |  |  |  |  |
| Regimen 1 | 86,360 | 9 | 0.101 | 0.886 | 0.652–1.20 | 0.440 |
| Regimen 2 | 86,618 | 10 | 0.115 | 1 | Reference |  |

CI, confidence interval; IMV/ECMO, invasive mechanical ventilation/extracorporeal membrane oxygenation

Figure 1 shows the daily cumulative probability of presenting primary and secondary outcomes.

**Figure 1.**
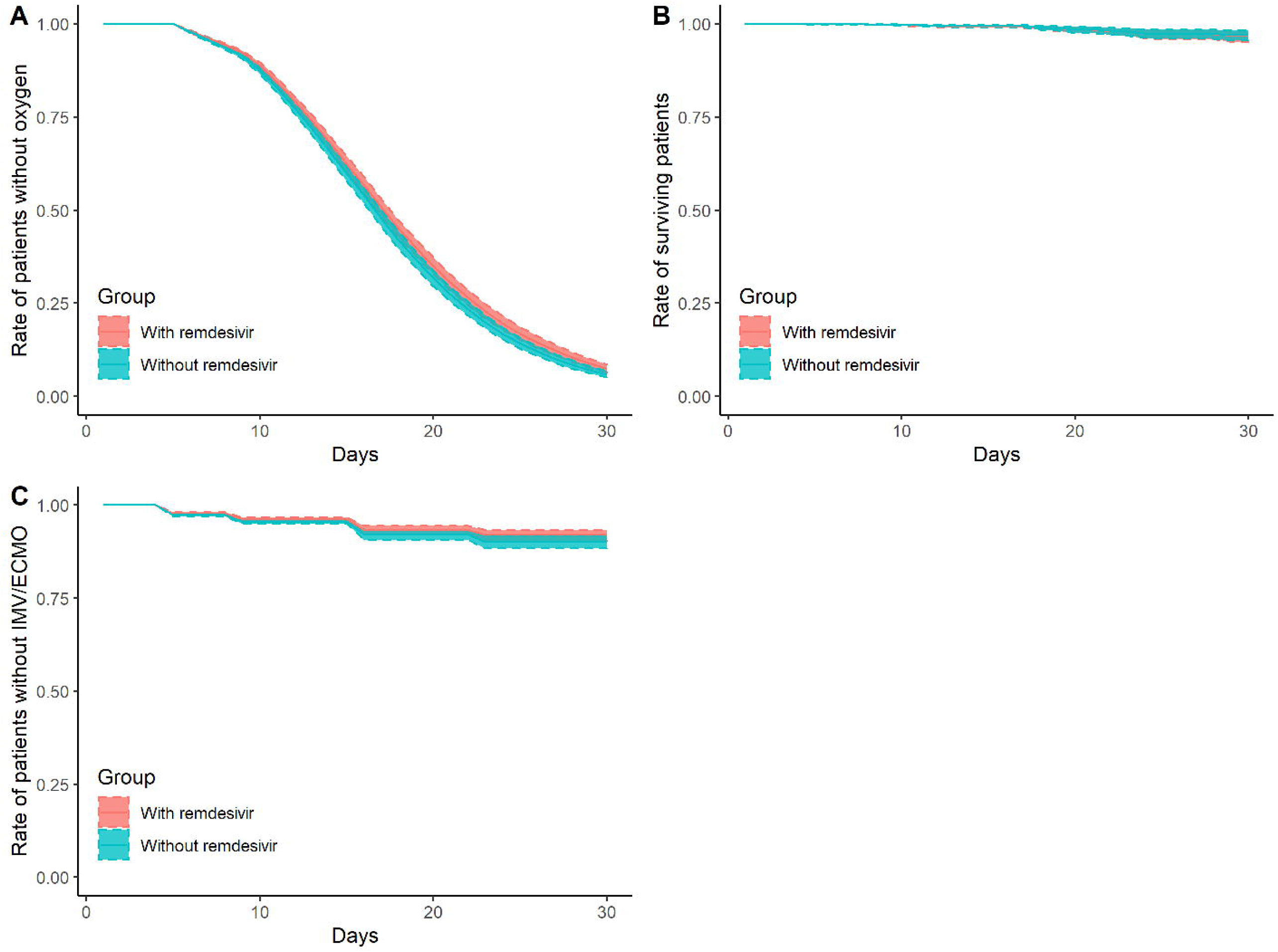
Daily cumulative probability of presenting primary/secondary outcomes. Panel A: daily cumulative probability of not being supported by oxygen Panel B: daily probability of survival Panel C: daily cumulative probability of not being supported by invasive mechanical ventilation (IMV)/extracorporeal membrane oxygenation (ECMO) Red ribbons represent Regimen 1 (treated with remdesivir) and blue ribbons represent Regimen 2 (treated without remdesivir). Shaded zones represent pointwise 95% confidence intervals by bootstrapping.

Regarding the safety of remdesivir treatment, 92 of 828 (11.1%) cases reported adverse events (Table 3), of whom 24 (26.1%) were considered as having probable relevance to remdesivir. Although 66 patients (71.7%) continued remdesivir treatment despite adverse events, the remaining 26 (28.3%) suspended their treatment. A total of 45 patients (48.9%) had liver dysfunction or liver enzyme elevation, 10 (10.9%) reported renal dysfunction, 3 (3.3%) had nausea/vomit, and 4 (4.3%) showed rash. No patient had sequelae due to adverse events.

**Table 3.**
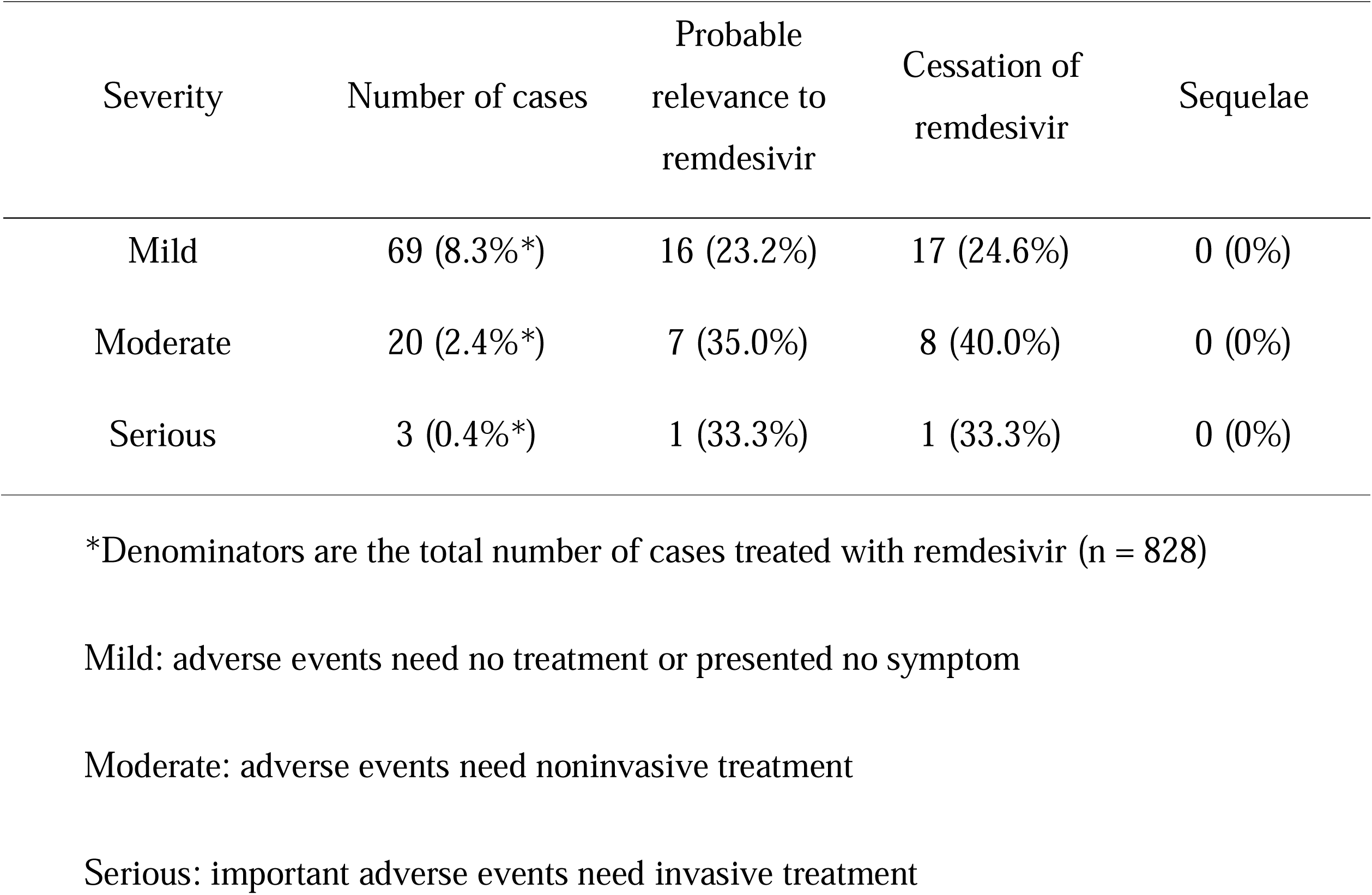
Adverse events during remdesivir treatment.

## Discussion

Our study showed that remdesivir administration in the early stage of disease might reduce supplementary oxygen requirement during hospitalisation. However, it did not reduce fatality risk and risk of IMV/ECMO requirement in hospitalised COVID-19 patients. These results concerning fatality and IMV/ECMO are compatible with a previous study[6] and support that remdesivir is not an essential drug for COVID-19-specific treatment, as suggested by the latest clinical guideline[8,24,25]. Similarly, the present study demonstrated the possible benefit of using remdesivir in the early stage of the disease. Our results suggest that among nonsevere hospitalised patients (i.e., patients without oxygen requirement), early initiation of remdesivir is beneficial.

However, further discussion would be desirable because our analysis demonstrated that the drug of interest did not improve the final prognosis of the disease. Although the risk of severe adverse events due to remdesivir appears to be low, we can consider it as an unnecessary risk if the drug does not improve the outcome. Conversely, substantial benefit can be obtained by reducing the burden on healthcare facilities if it prevents COVID-19 patients from the need for supplementary oxygen therapy.

Furthermore, we must note the fact that the healthcare system in Japan allowed us to hospitalise even nonsevere patients. For instance, Japanese indications for hospitalisation are quite different from those of other countries[8,25,26]; therefore, it is difficult to apply our results directly to different settings. In addition, the hospitalisation criteria in Japan have been changing over the COVID-19 pandemic time[27]. Initially, the indication of remdesivir in Japan was limited to severe COVID-19 cases[28]. Remdesivir was approved in Japan in May 2020 by the fast-track approval followed by the US FDA Emergency Use Authorization[29]. At that time, the indication of remdesivir was limited to severe patients whose oxygen saturation was ≤94% (ambient air) and who required supplementary oxygen or IMV/ECMO. In January 2021, the Ministry of Health, Labour and Welfare in Japan extended its indication to ‘patients who have pneumonia due to SARS-CoV-2 infection.’ Consequently, the number of mild or moderate cases administered remdesivir increased recently, and it enabled us to analyse the efficacy of remdesivir in the early stage of disease. Further evaluation in other healthcare settings will be one of the future challenges.

Our study has some limitations. The most important one is that it is not an RCT but a retrospective cohort study. Certainly, we sincerely attempted to adjust various factors that affect clinical outcomes; however, our method does not enable us to adjust all the numerous confounding factors[30]. Although our method enables us to adjust time-dependent factors and immortal time bias[13,14], we could not include time-dependent variables other than NEWS. Moreover, as our data are based on a registry system, it is difficult to interpret several items. For instance, ‘fatality’ in this study implies that a patient died during his/her 30-day observation period, i.e., during hospitalisation. Even if a patient died after he/she was discharged, we labelled this patient as a survived one. The cause of death is also not available from the registry data, and when a fatal case has a serious comorbidity such as cancer, we are not aware of the disease that caused death to the patient. Furthermore, COVIREGI-JP does not collect information about the daily clinical status of each patient. The adverse events of remdesivir were reported based on researchers’ decisions and thus might be underreported. Nevertheless, our data at least appropriately adjust the time-independent characteristics of patients associated with clinical outcomes (e.g., age, comorbidity, etc.) and an important time-dependent factor deeply associated with their clinical course and outcome (i.e., NEWS); hence, we believe that the results were reliable.

## Conclusions

Remdesivir might reduce supplementary oxygen requirement during hospitalisation. However, it showed no positive effect on the clinical outcome.

## Data Availability

Data used in this study would be available upon a reasonable request to the corresponding author.

## Acknowledgements

We thank all the participating facilities for their care of COVID-19 patients and cooperation in data entry.

## Ethics

This study was approved by the NCGM ethics review (NCGM-G-003494-0). Information regarding opting out of our study is available on the registry website.

